# The Blood-Brain Barrier in Bipolar Disorders: A Systematic Review

**DOI:** 10.1101/2022.11.08.22281826

**Authors:** Clara Wakonigg Alonso, Frances McElhatton, Brian O’Mahony, Thomas Pollak, Paul R.A. Stokes

## Abstract

**Background:** Bipolar disorders (BD) are chronic, debilitating disorders. Our understanding of the pathogenesis and functional biomarkers in BD remains limited. The blood-brain barrier (BBB), a highly selective, protective physical barrier which separates the central nervous system from the peripheral circulation, has been increasingly investigated in the BD. This systematic review aimed to assess the relationship between BD and markers of BBB dysfunction.

**Methods:** Studies were identified in PubMed and Medline databases in January 2021. Articles were limited to full-length peer-reviewed journal publications with no date restrictions. Included studies compared blood, CSF, post-mortem, genetic and imaging measures of BBB function in people with BD compared to healthy controls.

**Results:** 49 studies were identified, 34 of which found an association between BD and markers of BBB dysfunction. Blood QAlb, S100B and MMP levels were found to be increased in BD participants compared to controls in 57% of the studies. In post-mortem BD studies, ICAM, neurexin, claudin-5, and chondroitin sulphate proteoglycans were increased in the anterior cingulate cortex (ACC), prefrontal grey matter, occipital cortex and cerebellum, and lateral nucleus of the entorhinal cortex respectively compared to controls. Additionally, a study of BBB leakage measured by MRI found that nearly 30% of BD participants had extensive BBB leakage compared to controls. The mood state of BD participants was also associated with markers of BBB dysfunction, with participants experiencing mania generally having increased BBB marker levels compared to participants who were depressed or in remission.

**Conclusions:** This review suggests an association between BD and markers of BBB dysfunction. Further research is needed to control for a number of confounding factors, and to clarify whether this association provides a pathogenic mechanism, or is an epiphenomenon of BD.

## 1. Introduction

Bipolar disorders (BD) are chronic and highly debilitating psychiatric disorders which affect an estimated 45 million people worldwide (James et al., 2018). The aetiology of BD is still poorly understood but is conceptualised as a multifactorial process in which the environment and genetics play a role (Rowland & Marwaha, 2018). The involvement of the blood-brain barrier (BBB), a highly selective, protective physical barrier that separates the central nervous system (CNS) from the peripheral circulation, has been increasingly investigated in the aetiology of BD. The BBB is critical to ensure a controlled environment within the CNS (Abbott, Rönnbäck & Hansson, 2006), maintaining a specific ionic environment that favours appropriate neuronal activity and shields the brain tissue from toxic materials carried in the circulation (Abbott, 2010) (see Figure 1A).

**Figure 1.**
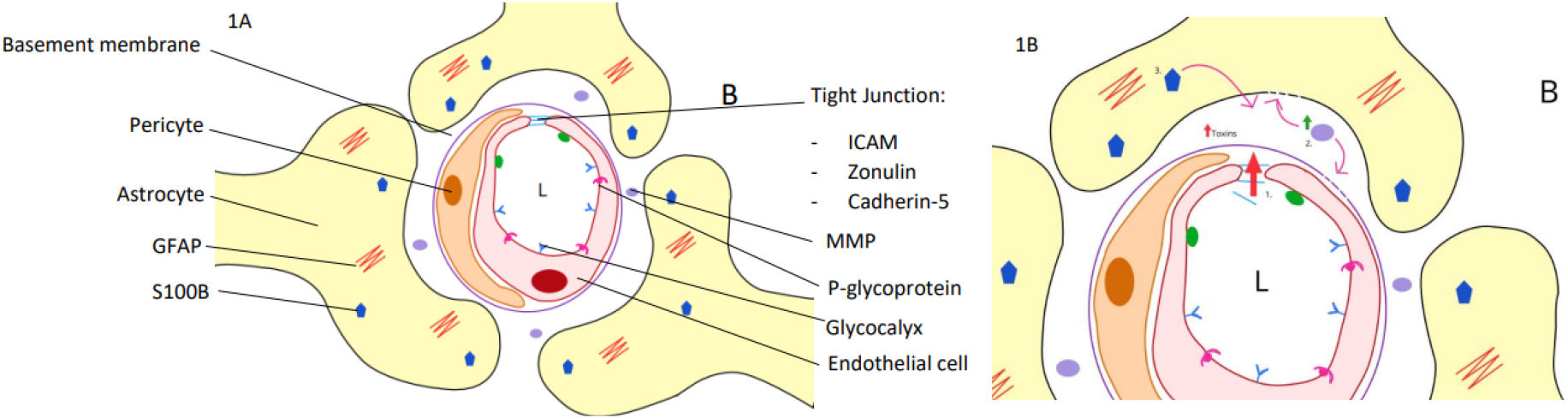
Blood-Brain Barrier structure (Fig 1A) and the potential effects of its breakdown (Fig 1B) 1A. Single cell endothelial cells form the lumen of the blood vessel. These are joined by tight junction and cell adhesion proteins. A basement membrane covers these endothelial cells. Pericytes sit around this structure without covering them entirely. Astrocyte end-feet cover the whole structure. GFAP glial fibrillary acidic protein, S100B calcium-binding protein B, MMP matrix metalloproteinase. B brain, L lumen 1B. A model of potential BBB dysfunction in BD. Breakdown of tight junction proteins (ICAM, Zonulin and Cadherin-5), may lead to increased diffusion of proteins and toxins from the bloodstream into the brain tissue. Increased toxin concentration may cause MMPs to activate and excessive MMP activation may be associated with basal membrane and astrocytic damage. S100B secreted from astrocytes may diffuse into the blood due to BBB dysfunction. BBB dysfunction may lead to a neuroinflammatory response, which in turn affects other cerebral tissue, such as neurons and their functioning.

Recent models propose that dysfunction of the BBB leads to increased permeability and reduced protection of the brain, allowing peripheral damaging substances into the cerebral tissues. This, in turn, may activate immune responses, such as microglial activation, and inflammatory responses which can disrupt healthy brain function (Patel & Frey, 2015) (see Figure 1B).

Although there is well-established evidence for altered BBB function in many neurological disorders, such as epilepsy, multiple sclerosis, and dementia (Gorter, Van Vliet, and Aronica, 2015) (Pinheiro, 2016) (Burgmans, Van de Haar, Verhey, and Backes, 2013), the role of BBB dysfunction has not been as extensively researched in psychiatric disorders. Pollak et al. (2018) reviewed the relation between BBB integrity and psychosis, concluding that disruption in the BBB could be contributing to psychosis due to neuronal and synaptic dysfunction, increased permeability into brain tissue and disrupted glutamate homeostasis. However there has not been a recent review that has examined the role of the BBB dysfunction in BD. This systematic review aimed to identify and synthesise relevant studies which have investigated the role of the BBB in BD.

## 2. Methods

### 2.1. Design

This study was a narrative systematic review conducted according to the Preferred Reporting Items for Systematic Reviews and Meta-Analyses (PRISMA) guidelines (Moher, Liberati, Tetzlaff, & Altman, 2009). The study protocol was registered with the UK National Institute for Health Research PROSPERO international prospective register of systematic reviews (ID, CRD42021228324). The review sought to include studies which measured BBB structure and function in people with BD and that compared findings to a healthy participant comparator group.

### 2.2. Included studies

Studies were included if they assessed BBB function or structure in participants of any age or sex, who met ICD or DSM criteria for BD (including BD type I, type II and not otherwise specified) and compared findings to a matched healthy participant control group. Only studies written in English were included. Studies which included participants with a diagnosis for cyclothymia and/or dysthymia, and animal studies were excluded from the review.

### 2.3. Outcome measures

Markers of BBB structure or function in people with BD compared to healthy participants was the primary outcome measure. BBB status was measured using a) blood- and CSF-based measures such as the CSF:serum albumin ratio (QAlb), lymphocyte concentration, serum concentration of brain-blood barrier associated proteins, b) post-mortem histological assays, c) neuroimaging studies, and d) studies of BBB-relevant genetic polymorphisms.

### 2.4. Data sources and searches

A systematic literature search for published studies was performed using PubMed, MEDLINE, PsycINFO and EMBASE from inception until 26/01/2021. The search was conducted by two independent reviewers following the same protocol, included studies were compared and disputes resolved by consensus. All studies included in the review were evaluated against pre-defined inclusion criteria by two of the review authors (CW and FM). Any disparities were addressed by reaching agreement via an additional review author (PS). No disagreements came up between the reviewers. The full list of search terms can be found in the appendix.

### 2.5. Data extraction

For each study, the following information was extracted where available: study type, sample size, intervention components, type of BD, mood state during sample collection, medications used, whether BD participants were in/outpatients, measure of BBB function and outcomes. Only data relating to BD and healthy control groups were extracted.

### 2.6. Quality assessment

The Newcastle-Ottawa scale was used to evaluate the quality of included case-control studies (Wells et al., 2021) and the Joanna Briggs Institute (JBI) Checklist for Analytical Cross-Sectional Studies (Joanna Briggs Institute, 2017) was used to evaluate cross-sectional studies. Risk of bias tables indicating the quality of included studies were created. See supplementary tables 1 and 2, respectively.

## 3. Results

### 3.1. Study Selection and characteristics

The study selection process is shown in PRISMA flow chart (see Figure 2).

**Figure 2.**
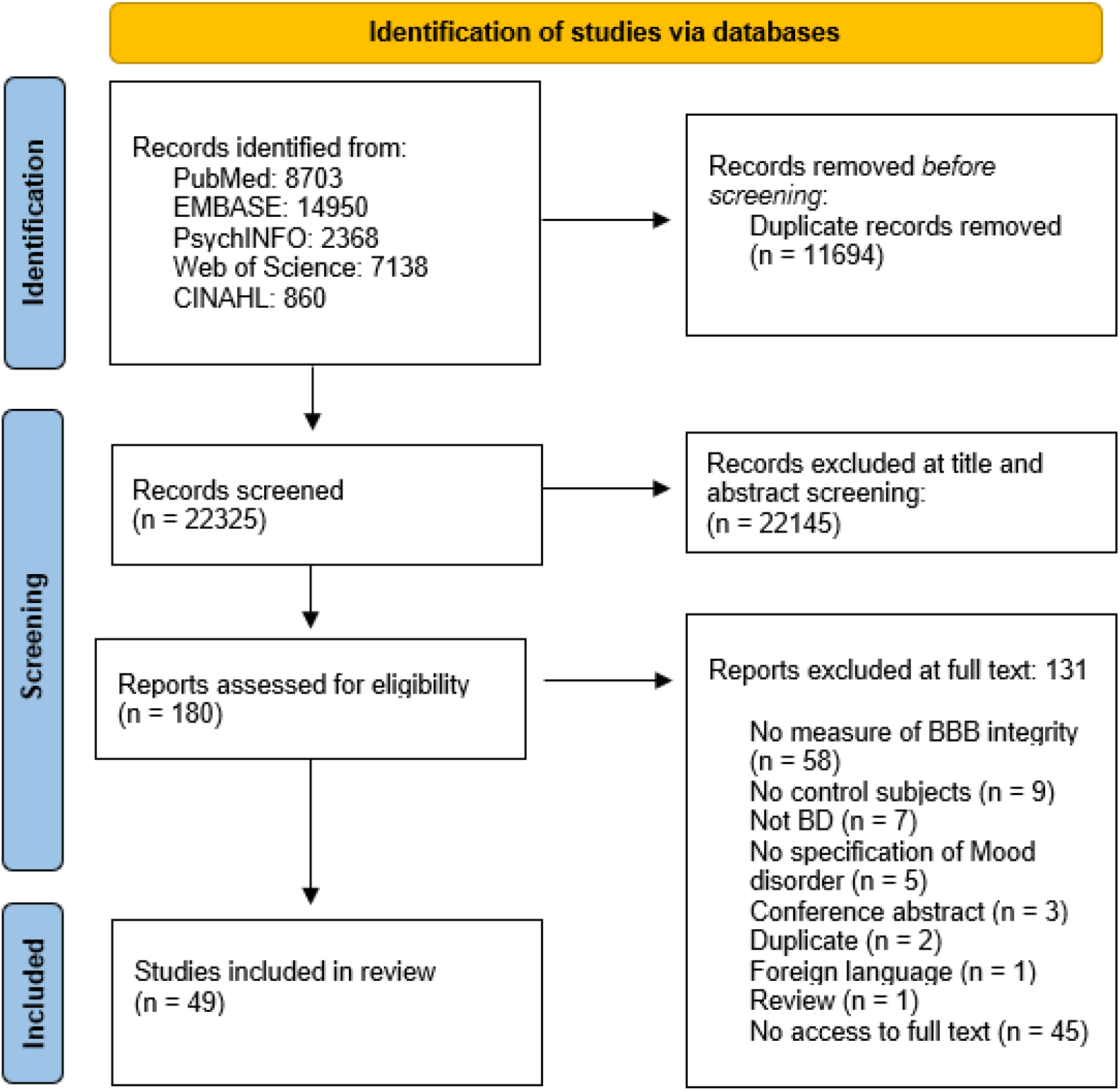
PRISMA flowchart summarising literature search and study selection

Supplementary Table 1 shows a summary of the characteristics of the studies included in this systematic review. All included studies were observational. 23 studies investigated blood/CSF markers, 19 studies used a post-mortem methodology, 7 used a genetic approach and one study used magnetic resonance imaging (MRI) to assess the integrity of the BBB. The studies included heterogenous samples of people with BD in terms of illness duration, severity, subtype, current affective state, and past and current medication use. Most BD participants were taking psychotropic medication at the time of the study.

### 3.2. Blood/CSF biomarker results

17 out of 23 studies included reported an association between blood or CSF markers of BBB dysfunction in BD; 13 found a difference between BD and healthy controls and 7 found a difference between people with a diagnosis of BD I vs BD II, or between BD manic/depression and remission groups (See Table 1).

**Table 1.**
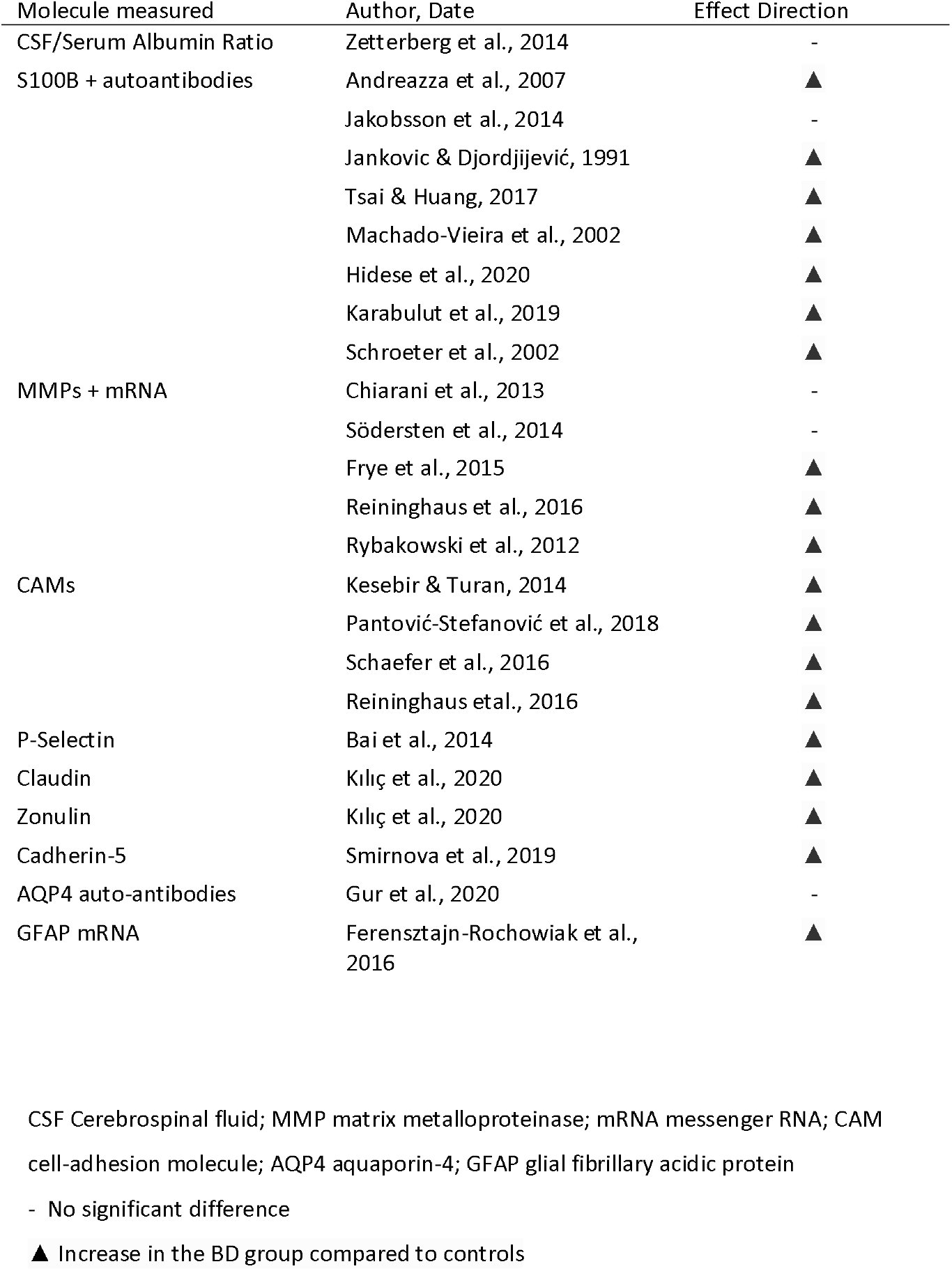
Summary of the association between bipolar disorder and Blood and CSF markers of BBB function

#### 3.2.1. QAlb

QAlb (CSF:serum albumin quotient) is the ratio of cerebrospinal fluid albumin to serum albumin. As albumin is exclusively synthesised in the liver, and in the healthy BBB is transported into the CNS only by passive diffusion, an increased QAlb indicates increased BBB permeability (Zetterberg, Mattsson & Blennow, 2010). Only one study measured the QAlb in BD participants compared to healthy participants. Zetterberg et al. (2014) found QAlb was significantly higher in all BD groups (BD I, II and NOS) compared to controls (*p*=0.017). Additionally, participants with BD I were found to have an elevated QAlb compared to participants with BD II (p<0.041). QAlb levels also correlated with lifetime episodes of psychosis (*p*=0.026) and was significantly increased in participants currently prescribed antipsychotic medication. When antipsychotic medication history was adjusted for, diagnosis and lifetime psychosis no longer had a significant effect. No difference in QAlb was found between BD participants not currently prescribed antipsychotics and healthy participants.

#### 3.2.2. S100B

S100B is a protein which is predominantly synthesised and secreted by astrocytes. The normal concentration of S100B in blood is generally low and an increase in S100B serum concentration may indicate reduced BBB integrity (Strathmann, Schulte, Goerl & Petron, 2014) (Arora et al., 2019). The presence of S100B antibodies has also been suggested to indicate a long-term reduction in BBB integrity (Marchi et al., 2013; Choi et al., 2016). 8 studies measured S100B levels in people with BD and in healthy controls (see Table 1). Results of the blood-based S100B studies were mixed: Machado-Vieira et al. (2002) and Schroeter et al. (2002) compared participants with mania to age-matched healthy participants and found that S100B levels was significantly higher in mania (p=0.014). Andreazza et al. (2007) found S100B levels to be significantly higher in mania (*p* = 0.011) and bipolar depression (*p* = 0.004) but was not altered in remission. Tsai & Huang (2017) reported no significant difference in S100B levels between BD participants and healthy controls but did find a significant reduction in S100B levels between BD participants when in a manic episode compared to remission. Karabulut et al. (2019) found that S100B levels were higher in participants with BD with a longer duration of illness. Neither of the S100B CSF studies (Jakobsson et al., 2014 and Hidese et al., 2020) found any significant difference between BD and healthy groups. Jankovic & Djordjijević (1991) found that significantly more participants with BD (28.2%) had anti-S100B antibodies than controls (2.7%).

#### 3.2.3. MMPs

Matrix metalloproteinases (MMPs) are enzymes that activate as a result of vascular endothelial damage and can lead to degradation of the extracellular matrix, including the proteins that form tight junctions (Van den Steen et al., 2002; Lischper et al., 2010; Feng et al., 2011). An increase in MMP concentration can potentially represent both a marker and a cause of BBB breakdown. Studies comparing MMP levels and/or expression in BD reported mixed results: 3 reported increased MMP levels in participants with BD compared to healthy controls (Frye et al.2015; Reininghaus et al., 2016; Rybakowski et al., 2012) while two found no differences (Chiarani et al., 2013; Södersten et al., 2014). Among the significant associations Frye et al. (2015) found increased MMP-7 levels in BD compared to controls (p=0.009), and also found increased levels in BD I participants compared to BDII. Reininghaus et al. (2016) found that serum MMP-9 was increased in those with a longer duration of illness (p = 0.006).

#### 3.2.4. ICAM and VCAM

ICAM and VCAM are cell adhesion molecules which are involved in the regulation of leukocyte transport across the BBB. The endothelial cells that form the BBB upregulate expression of ICAM and VCAM in response to inflammation, thus increasing BBB permeability to leukocytes (Daneman, & Prat, 2015). Four studies measured the serum concentration of cell adhesion molecules, and each reported an association between VCAM/ICAM levels and BD (Kesebir & Turan, 2014; Pantović-Stefanović et al., 2018; Schäfer et al., 2016; Reininghaus et al., 2016). Schäfer et al. (2016) found that ICAM was significantly increased in people with BD compared to controls regardless of the type of BD or the participant’s mood state, with a greater increase in manic vs depressed patients. Reininghaus et al. (2016) found that overall, ICAM concentrations were increased in all BD groups studied (p = 0.006) and that BD participants with a longer duration of illness had higher ICAM concentrations compared to those with a more recent diagnosis. Pantović-Stefanović et al. (2018) reported elevated ICAM levels in BD *(p*□=□0.021), while also finding that people with BD had lower VCAM levels (*p*□=□0.000). Kesebir & Turan (2014) investigated ICAM and VCAM levels in participants with a first manic episode and found that both markers were significantly elevated in comparison to the same participants during subsequent remission, as well as compared to healthy controls (*p-values* <0.001).

#### 3.2.5. Other markers

Several studies have investigated other potential markers of BBB integrity in BD. Soluble P-selectin receptor (sP-selectin) has been implicated in contributing to BBB dysfunction (Kisucka et al., 2009; Jin et al., 2010) and Bai et al. (2014) found higher sP-selectin concentrations in BD (*p*=0.029). Kılıç et al. (2020) found that claudin and zonulin, both components of the tight junctions between the endothelial cells of the BBB (Sturgeon & Fasano, 2016; Greene, Hanley & Campbell, 2019) were elevated in BD I during mania and in remission compared to controls (*p* < .001). Smirnova et al. (2019) reported no difference between cadherin 5 serum levels, a transmembrane protein that forms the adherens junctions between the endothelial cells that form the BBB (Kuriyama et al., 2014), in BD compared to controls (p=0.45).

#### 3.2.6. Effect of mood state on BBB Blood/CSF markers

11 studies compared BBB dysfunction markers in BD participants in different mood states. 6 studies reported that markers of BBB dysfunction were significantly increased during manic or bipolar depressive episodes compared to euthymia (Andreazza et al., 2007; Tsai & Huang, 2017; Hidese et al., 2020; Kesebir & Turan, 2014; Rybakowski et al., 2012; Pantović-Stefanović et al., 2018). Hidese et al. (2020) found that CSF S100B levels were positively correlated with the severity of depression or mania in BD. S100B was found to be increased in manic episodes compared to depressive and euthymic episodes (Andreazza et al., 2007; Tsai & Huang, 2017; Hidese et al., 2020). Increased ICAM levels were also reported in manic episodes compared to remission as well as VCAM levels (*p*=0.049) (Kesebir & Turan, 2014). However, Pantović-Stefanović et al. (2018) found decreased levels in VCAM during mania compared to depression in BD (*p*□=□0.016). MMP-9 levels were found to be increased during mania and depression compared to remission (Rybakowski et al., 2012).See Table 2.

**Table 2.** Vote counting table showing direction of effects of mood state on BBB permeability markers

| Author, Date | Effect Direction | Molecule measured |
| --- | --- | --- |
| Andreazza et al., 2007 | ▲ | S100B |
| Tsai & Huang, 2017 | ▲ | S100B |
| Hidese et al., 2020 | ▲ | S100B |
| Karabulut et al., 2019 | - | S100B |
| Chiarani et al., 2013 | - | MMP-9 |
| Rybakowski et al., 2012 | ▲ | MMP-9 |
| Kesebir & Turan, 2014 | ▲ | ICAM & VCAM |
| Pantovic-Stefanovic et al., 2018 | ▲ for VCAM<br>- for ICAM | ICAM & VCAM |
| Schäfer et al., 2016 | - | ICAM |
| Bai et al., 2014 | - | P-selectin |
| Kiliç et al., 2020 | - | Zonulin & claudin |

MMP-9 matrix metalloproteinase-9; ICAM intercellular adhesion molecule; VCAM vascular cell adhesion molecule
- No significant difference
▲ Increase in the BD group compared to controls

### 3.3. Imaging studies

One study, Kamintsky et al. (2020), used dynamic gadolinium contrast-enhanced MRI to create quantitative maps of brain leakage, on the basis of which 50 participants (36 BD and 14 controls) were divided into extensive leakage and non-leakage groups. All 10 of the participants in the extensive BBB leakage group were people with BD, and all had insulin resistance. These BD participants displayed a diffuse, rather than a local, pattern of leakage across brain regions.

### 3.4. Post-mortem studies

All 19 studies (which included 889 participants) used q-PCR and/or immunostaining for the quantification of BBB markers including astrocyte markers (GFAP, S100B and ALDH1L1), tight junction proteins (Claudin-5), cell adhesion proteins (ICAM, VAM and neurexin) and glycocalyx proteins (chondroitin sulphate proteoglycans). All the extracted analyses were from people with BD compared to controls. GFAP studies were not included in this review due to its unreliability as a BBB marker (Sofroniew & Vinters, 2009). A summary of the results can be found in Table 3.

**Table 3.**
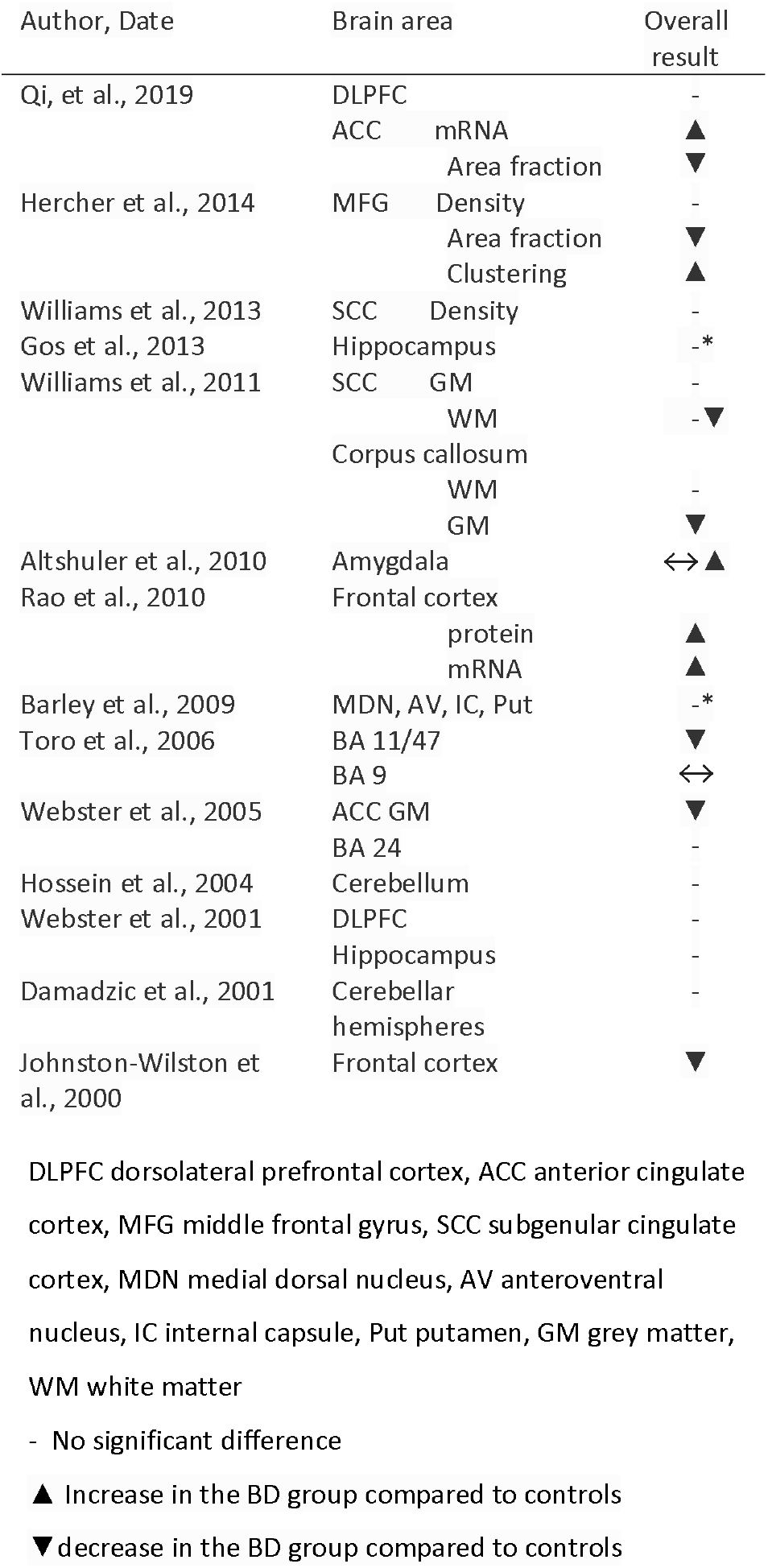
GFAP post-mortem studies: results summary.

Cell adhesion molecules (ICAM and neurexin) were reported to be increased in the ACC (p=0.001) and the prefrontal grey matter (p=0.01) in people with BD compared to controls (Jenkins et al., 2016) (Thomas et al., 2016). Greene et al., (2020) reported increased Claudin-5 mRNA levels in the occipital cortex (p=0.05) and cerebellum in people with BD (p=0.03) which also correlated with suicide and age. Chondroitin sulphate proteoglycans levels were increased in the lateral nucleus of the entorhinal cortex (EC) (p=0.04), decreased in layer III of the EC (p=0.02) as well as in the amygdala (p<0.03) in BD (Pantazopoulos et al., 2010 and 2015) and this also correlated with lithium medication history. See Table 4.

**Table 4.** Tight junctions and cell adhesion molecule post-mortem studies: results summary.

| Author, Date | Brain region | Overall results |
| --- | --- | --- |
| Greene et al., 2020 | Hippocampus vessel density | -<br>▲ |
|  | OFC vessel diameter | - |
|  | Claudin 5 levels | ▲ |
|  | Occipital cortex and cerebellum Claudin-5 mRNA |  |
| Jenkins et al., 2016 | PFC GM neurexin levels | ▲ |
| Pantazopoulos et al., 2015 | Amygdala aggrecan glial cells | -<br>▼ |
|  | 3B3IR glial clusters |  |
| Pantazopoulos et al., 2010 | EC CSPG LN | ▲ |
|  | ECLIII | ▼ |
| Thomas et al., 2004 | ACC ICAM GM | ▲ |
|  | WM | ▲ |
|  | DLPFC ICAM | - |

OFC orbitofrontal cortex, PFC prefrontal cortex, EC entorhinal cortex, LN lateral nucleus, ACC anterior cingulate cortex, DLPFC dorsolateral prefrontal cortex
- No significant difference
▲ Increase in the BD group compared to controls
▼ decrease in the BD group compared to controls

A full description of the results available in Supplementary Table 3.

### 3.5. Genetic studies

A handful of studies looked at BBB-relevant genetic loci in relation to medication response in BD. Rybakowski et al. (2011) studied lithium response in BD in relation to the genotype coding for the MMP9 protein and found no significant deviation from equilibrium in the distribution of the polymorphisms. P-glycoprotein and MMP-9 polymorphisms have been explored in other studies. Rybakowski et al., (2009) found that the functional polymorphism (rs3918242) of the MMP9 gene to be more common in BD than controls (p=0.02). This was not the case for the MMP3 functional polymorphisms (Kucukali et al., 2009). For the P-glycoprotein polymorphisms, Turgut et al. (2009) found a deviation from equilibrium in the C3435T polymorphism, indicating a possible association between BD and p-glycoprotein structure (p<0.01). Naumovska et al. (2017) found an uneven distribution of p-glycoprotein polymorphisms only in women, with the alleles 1236 and 2677 being potential risk polymorphisms for BD in women (p<0.05). In the case of cell adhesion molecule genes, 22 significant risk genes were identified (See Table 5). No associations were found to other factors apart from sex. More information on these studies is presented in Supplementary Table 3.

**Table 5.** BBB genetic markers results summary

| Author, Date | Gene | Overall result |
| --- | --- | --- |
| Naumovska et al.,<br>2017 | ABCB1 overall<br>SNP 1236 and 2677 W vs<br>M | -<br>? |
| Rybakowski et al.,<br>2011 | MMP9 | - |
| Kuckali et al., 2009 | MMP3 | - |
| Rybakowski et al.,<br>2009 | MMP9 | ? in T allele |
| Turgut et al., 2009 | MDR1 | ? in CT<br>genotype |
| De Luca et al., 2003 | MDR1 | - |
| O'Dushlaine et al.,<br>2011 | CAM<br>TJ | 22?*<br>16?* |

? differential distribution between bipolar and controls. Specific genotype or allele increased in bipolar subjects. - no significant deviation
\* Number denotes the amount of different genes which were differentially distributed in BD subjects
SNP single-nucleotide polymorphism

## 4. Discussion

The results of this systematic review provide converging evidence to suggest the involvement of BBB dysfunction in BD. While a synthesis of the literature is challenging given the heterogeneity in study methods, participants and design, there is some evidence for BBB dysfunction from serum-based and CSF-based, post-mortem, neuroimaging, and genetic studies. The most extensive and perhaps convincing evidence is from serum and CSF studies, where 17 out of 23 included studies found evidence in favour of an association between BD and markers of BBB disruption, including 13 studies reporting a difference in levels of BBB markers between participants with BD and healthy controls, and six studies reporting a difference in levels of BBB markers depending on the mood state of participants with BD.

Measurement of BBB permeability in the living human brain is challenging and perhaps for this reason most studies in this review include a proxy marker of the state of the BBB. Perhaps the most direct measures of permeability are QAlb and the radiographic analysis of intracerebral leakiness. Only one study of each of these measures in BD has been published, and while both reported increased BBB permeability in BD to controls, it is clear that further studies of each type are required. The paucity of QAlb studies is particuarly surprising, given how numerous similar studies are in the schizophrenia/psychosis literature (Pollak et al., 2018). Overall, studies of serum and CSF markers of BBB function in BD included in this review are at best suggestive of dysfunction. One reason for this is that in most cases BBB-specific cell types are not the only source of the measured proteins and so these findings could also be measuring damage to other cell types (e.g., S100B and astrocytic damage). Nonetheless, these studies provide important mechanistic leads.

From our review, the evidence for BBB dysfunction being associated with specific mood states in BD was unclear. Of the blood/CSF studies, only 11 of the 23 studies performed subgroup analysis between different mood states and the results were mixed. 6 found that markers of BBB permeability were increased during acute episodes of depression or mania compared to euthymia and/or healthy controls or that the level of BBB permeability markers was associated with the severity of the acute episode. This may suggest that BBB permeability is involved in the pathophysiology of acute episodes but may also represent a transient phenomenon. The association between mood states in BD and BBB dysfunction remains a highly promising focus of future research.

Given the mechanistic links between BBB dysfunction and neuroinflammation, the suggestion that there may be state-specific effects is consistent with an emerging literature suggesting state-specific increases in various inflammatory markers in acute mood states in BD (Leboyer et al., 2012; Dickerson et al., 2013). Two studies, Kamintsky et al. (2020) and Karabulut et al. (2019), found a positive correlation between the chronicity of BD and measures of BBB permeability. This might suggest that BBB permeability increases with, and may be a marker of, chronicity or progression of disease, while also representing a confounding factor for past and future studies. Carefully controlled work comparing people with BD with different degrees of disease progression across multiple BBB biomarkers will be necessary to determine whether BBB dysfunction increases with disease progression.

The heterogeneity of findings and use of indirect markers of BBB dysfunction likely means that no clear conclusion can be drawn from the included post-mortem studies of BBB markers in BD. The results included in this review indicate variation of BBB permeability throughout the brain, rather than a consistently increased permeability in any particular brain area. Cell adhesion molecules, such as ICAM, are expressed in endothelial cells of the vasculature and may be deregulated by inflammation (Dietrich, 2002). The increased levels reported by the studies included in this review could indicate an inflammatory response in the brain with upregulation of ICAM in the brain vasculature rather than a breakdown of the BBB. The genotype studies we included also identified a significant association with single-nucleotide polymorphisms (SNPs) in the genes coding for the proteins of the cell adhesion and extracellular matrix pathways in people with BD (O’Dushlaine et al., 2011) which mirrors findings in schizophrenia (Zhang et al., 2015; Rempe, Hartz, & Bauer, 2016).

The effect of medication on levels of markers of BBB dysfunction was investigated by 26 studies included in this review. 18 studies found no effect of medication on markers of BBB integrity, regardless of the type of medication examined. S100B levels were reported to be lower in BD participants who were taking psychotropic medications, however the specific medications used were not indicated (Hidese, 2020; Tsai & Huang., 2017; Schoeter et al., 2002). QAlb and GFAP were reported higher in BD participants treated with antipsychotics (Zetterberg et al., 2014; Toro et al., 2006). Lithium treatment was positively correlated to increased glial cell levels (Pantazolopoulos et al., 2015; 2010). Given these findings, there is no conclusive evidence that pharmacotherapy in BD is associated with BBB dysfunction, but this is an area that requires further detailed investigation.

Overall, the quality of the included studies in this review was acceptable. Case-control studies assessed with the Newcastle-Ottawa scale found that all studies, except for two (O’Dushaine et al., 2011; Rybakowski et al., 2009), reached high scores, indicating a low probability of bias. Meanwhile, cross-sectional studies assessed using the Joanna Briggs Institute Checklist for Analytical Cross-Sectional Studies found that more than half of the studies had a high-quality marking, while ten were moderate and only one ranked as low quality (Jankovic & Djordjijević, 1991) (Supplementary tables 1 and 2).

## Limitations

There are several limitations to the methodologies used in the studies included in this review. Measuring post-mortem levels of BBB related proteins provides at best an indirect measure of BBB integrity. Imaging methods have been used to assess the integrity of the blood-brain barrier very successfully (Chassidim et al., 2013) however this has only been used in one study in BD. Not every CSF/serum marker explored in this review are well established as markers of BBB dysfunction. The two most robust markers are CSF/serum albumin ratio, considered the gold standard as a marker of BBB permeability, (Roh, Cho, Yoon, & So, 2017) and S100B measured in blood/CSF which has been demonstrated multiple times to be a valid marker of BBB damage (Marchi et al., 2004; Koh & Lee, 2014). This review only found one study that measured CSF/serum albumin ratio (Zetterberg et al., 2014). Importantly, because of the diversity of methodologies used in the studies included in this systematic review, heterogeneity between study protocols was not sufficient to allow a meta-analysis.

Additionally, reporting of BD participant characteristics was poor with some studies failing to report potentially relevant variables such as mood state at time of study, specific BD diagnosis and illness duration. Many studies did not conduct any sub-group analysis within BD groups based on these characteristics; particularly lacking were studies that considered BD progression/illness duration. Therefore, there were a limited number of studies on which to draw conclusions about the specific nature of the association between BD and BBB dysfunction which may limit the validity of the findings in this review. The included age range in the blood/CSF papers was also quite narrow, particularly excluding older adults which may have affected these results. Many of the post-mortem studies had small sample sizes, which could have affected the study power, potentially causing less robust effects to fall short of statistical significance.

### Future research

More research is needed to conclusively address the question of whether BBB dysfunction is associated with BD, particularly regarding associations with mood state. This review found significant limitations with the current literature, especially in the heterogeneity between studies. We would suggest that future studies should compare participants with different BD types (I, II, NOS, with and without psychosis) and in different mood states to explore the nature of the potential association of BBB dysfunction and BD. Medication use should also be controlled for and reported. Additionally, studies should classify BD participants by illness duration and aim to clarify whether BBB plays a role in BD progression. We would also suggest that studies with the objective of measuring BBB dysfunction should measure CSF/serum albumin ratio and serum S100B levels as these are currently the two most reliable markers of BBB dysfunction, however further research into the validity of other markers of BBB dysfunction is also necessary. Further larger neuroimaging studies of BBB integrity are also needed in BD as only one imaging study has been published and this provided promising preliminary results

## 5. Conclusions

This review provides evidence towards an association between BBB dysfunction, specifically increased BBB permeability, and bipolar disorders. It is unclear whether this relationship means that markers of BBB dysfunction may be a state or trait marker of bipolar disorders. Markers of blood-brain barrier dysfunction have been shown to be correlated with chronicity of bipolar disorder, which could be due to primary processes of the disorder or its treatment, worsening physical health, or other psychosocial factors. As such, it remains to be seen to what extent these pathologies co-occur, and in what proportion of people with bipolar disorders. Further research, ideally longitudinal follow-up studies, is needed to answer these questions.

## Supporting information

Appendix - search terms

## Data Availability

All data produced in the present study are available upon reasonable request to the authors

## 6. Acknowledgments

This paper represents independent research part funded by the National Institute for Health Research (NIHR) Maudsley Biomedical Research Centre at South London and Maudsley NHS Foundation Trust and King’s College London. The views expressed are those of the authors and not necessarily those of the NHS, the NIHR or the Department of Health and Social Care.

## 7. Declaration of interests

TP has received a speaking honorarium from Janssen for an educational lecture on an unrelated topic. PRAS reports non-financial support from Janssen Research and Development LLC, personal fees and non-financial support from Frontiers in Psychiatry, personal fees from Allergan and a grant from H Lundbeck, outside the submitted work.

This research did not receive any specific grant from funding agencies in the public, commercial, or not-for-profit sectors.

## Notes

### Clinical Protocols

https://www.crd.york.ac.uk/prospero/display_record.php?RecordID=228324

