## Appendix - search terms for "The Blood-Brain Barrier in Bipolar Disorders: A Systematic Review"

Example PubMED:

**#1:** Depression[Text Word] OR depression[MeSH Terms] OR "major depressive disorder"[Text Word] OR depressive disorder, major[MeSH Terms] OR ”bipolar disorder”[Text Word] OR bipolar disorder[MeSH Terms] OR mania OR manic symptoms

**#2:** "blood brain barrier"[Text Word] OR BBB[Text Word] OR blood-brain barrier[MeSH Terms] OR “blood cerebrospinal fluid barrier”[Text Word] OR BCSFB[Text Word] OR s100b[Text Word] OR “S100 Calcium Binding Protein beta Subunit”[MeSH Terms] OR albumin[Text Word] OR QAlb[Text Word] OR “Immunoglobulin G”[Text Word] OR “IgG”[Text Word] OR “Immunoglobulin G”[MeSH Terms] OR fibrinogen[Text Word] OR plasminogen[Text Word] OR "matrix metalloproteinase"[Text Word] OR matrix metalloproteinases[MeSH Terms] OR MMP[Text Word] OR “tissue inhibitor of metalloproteinase"[Text Word] OR tissue Inhibitor of metalloproteinases[MeSH Terms] OR TIMP[Text Word] OR “cell adhesion molecule”[Text Word] OR cell adhesion molecules[MeSH Terms] OR selectin[Text Word] OR ICAM[Text Word] OR VCAM[Text Word]  OR glycocalyx[Text Word] OR glycocalyx[MeSH Terms] OR syndecan[Text Word] OR "heparan sulfate"[Text Word] OR "chondroitin sulfate"[Text Word] OR hyaluronan[Text Word] OR "glial fibrillary acidic protein"[Text Word] OR "glial fibrillary acidic protein"[MeSH Terms] OR GFAP[Text Word] OR "amyloid beta"[Text Word] OR amyloid beta-peptides[MeSH Terms] OR Aβ[Text Word] OR “p-glycoprotein”[Text Word] OR ATP Binding Cassette Transporter, Subfamily B, Member 1[MeSH Terms] OR “choroid plexus”

**#3:** #1 AND #2
